# Performance and validation of an adaptable multiplex assay for detection of serologic response to SARS-CoV-2 infection or vaccination

**DOI:** 10.1101/2022.05.16.22275163

**Authors:** Grace Kenny, Riya Negi, Sophie O’Reilly, Alejandro Garcia-Leon, Dana Alalwan, Colette Marie Gaillard, Gurvin Saini, Rosana Inzitari, Eoin R. Feeney, Obada Yousif, Aoife Cotter, Eoghan de Barra, Corinna Sadlier, Fiona Crispie, Peter Doran, Virginie Gautier, Patrick WG Mallon, the All Ireland Infectious Diseases cohort study

## Abstract

Measurement of quantitative antibody responses are increasingly important in evaluating the immune response to infection and vaccination. In this study we describe the validation of a quantitative, multiplex serologic assay utilising an electrochemiluminescence platform, which measures IgG against the receptor binding domain (RBD), spike S1 and S2 subunits and nucleocapsid antigens of SARS-CoV-2. The assay displayed a sensitivity ranging from 73-91% and specificity from 90 to 96% in detecting previous infection with SARS-CoV-2 depending on antigenic target and time since infection, and this assay highly correlated with commercially available assays. The within-plate coefficient of variation ranged from 3.8-3.9% and the inter-plate coefficient of variation from 11-13% for each antigen.

## Background

Most individuals develop a serologic response following infection with, or vaccination against SARS-CoV-2 ^1,2^. Multiple sources of evidence now reinforce the role of antibodies in protecting against infection or severe disease^3,4^. However there is significant variability in antibody response between individuals^5^, and a threshold titre of antibody necessary to provide protective immunity has not been established. Quantitative assays can help better map kinetics of the immune response following infection or vaccination and guide the need for, and optimal timing of, further booster vaccine doses, as well as appropriately directing use of increasingly available immunotherapies.

Vaccines against SARS-CoV-2 currently licensed by the European Medicines Agency (EMA) all target the spike protein^6^. For vaccine-induced immune responses, anti-spike antibodies correlate best with viral neutralising capacity^7^, the gold standard *in vitro* correlate of protection for many vaccine preventable diseases^8,9^. In contrast, post infectious anti-SARS-CoV-2 immune responses are usually characterised by a broader antigenic repertoire, and can be differentiated from vaccine-mediated immunity by the presence of anti-nucleocapsid antibodies. Given the increasing population prevalence of infection with SARS-CoV-2, distinguishing infection versus vaccine induced immunity may be important in detecting “breakthrough” infections in vaccinated individuals and potentially guiding vaccine sparing strategies, given the enhanced serologic response to vaccination in individuals with prior COVID-19^10^.

A wide array of serologic assays have been developed since the emergence of the pandemic, which vary in antigenic target, analytical platform employed and assay performance. The vast majority of these are either qualitative or semi-quantitative. Of 85 serologic assays with current emergency use authorisation (EUA) from the FDA, only one is fully quantitative^11^, and this targets only the S1 subunit. While useful as diagnostic or public health tools, qualitative and semi quantitative assays are currently insufficient to accurately define an immune correlate of protection against either infection or development of severe COVID-19.

To address the need for a quantitative immunoassay that detects both anti spike and nucleocapsid antibodies we developed an electrochemiluminescence (ECL) based multiplex serologic assay. ECL technology offers advantages over the more commonly used enzyme linked immunosorbent assays (ELISA) or indirect fluorescent antibody assays (IFA) in the quantitative measurement of SARS-CoV-2 antibody responses, as ECL based techniques allow for a broad dynamic range of measurements and increased sensitivity ^12^.

## Methods

We conducted a series of experiments in the UCD Centre for Experimental Pathogen Host Research (CEPHR) to develop a new, multiplex immunoassay to detect antibodies to SARS-CoV-2 (CEPHR Assay) and tested this assay on a series of available biobanked samples.

### Study Participants

The All Ireland Infectious Diseases Cohort Study is a prospective, multicentre study that enrols individuals presenting with issues pertaining to infectious diseases in hospitals in Ireland. Individuals provide written, informed consent for collection of demographic and clinical variables as well as blood for biobanking on up to 5 occasions every 6 months. Plasma, derived from ethylenediamintetraacetic acid (EDTA) anticoagulated whole blood, was stored at −80°C until analysis. For this analysis we included either individuals with polymerase chain reaction (PCR)-confirmed COVID-19, documented vaccination with 2 doses of mRNA-1273, BNT162b2 or ChAdOx1-S, at least 14 days from the second dose, or individuals without COVID-19 or vaccination recruited with available biobanked plasma dated no later than July 2019, prior to the onset of the COVID-19 pandemic.

### CEPHR COVID19 Serology Assay protocol

MesoScale Diagnostics (MSD) U-PLEX technology offers quantification of multiple analytes using multi-spot plates and electrochemiluminescence detection. U-PLEX development packs (MSD, Rockville, MD, USA) consist of multi-well plates comprising 96 wells pre-printed with 6 spatially separated binding reagents specific to U-PLEX “linkers”. Biotinylated reagents can be coupled to individual linkers, which then bind to distinct spots in each well, allowing for the development of customisable multiplex assays. In this assay, biotinylated SARS-CoV-2 receptor binding domain (RBD), spike S1 sub unit, spike S2 subunit and nucleocapsid (N) (Sino Biological Inc, Germany) were diluted to 1.25ug/ml, 5ug/ml, 4ug/ml and 3ug/ml respectively in ChonBlock ELISA buffer (Chondrex Inc, Redmond, WA, USA). Each biotinylated antigen was incubated with a different MSD U-PLEX “linker” for 30 minutes. MSD U-PLEX stop solution, which stops the coupling reaction, was then added and the mixture incubated for a further 30 minutes. All linkers were combined to make a coating solution. 50ul of coating solution was added to each well and incubated at room temperature for one hour, allowing the individual reagent-coupled linkers to self-assemble to spatially separated spots in each well. Plates were then washed 3 times with phosphate buffered saline (PBS) and 0.05% tween (Bio Sciences Ltd, Ireland).

Plasma samples were diluted 1:1600 in ChonBlock ELISA buffer and 25ul of diluted plasma was added to each well, with each plasma sample run in duplicate. To construct standard curves, RBD, S1, S2 and N antibodies (all derived from the sequence of the Wuhan-Hu-1 reference strain, Sino biological Inc, Germany) were diluted to neat concentrations of 6.25 ng/ml, 21 ng/ml, 37.5 ng/ml and 50 ng/ml respectively in ChonBlock ELISA buffer. These neat concentrations then underwent seven serial dilutions (1:2) to make a seven point standard curve, with 25ul of standard added per well. Each plate included two positive controls; a high and low titre plasma sample from individuals recovered from COVID-19. The plate containing the samples and standard curves were then incubated for 30 minutes at room temperature, washed, following which MSD SULFO-TAG-labelled goat anti-human IgG secondary antibody was added at a concentration of 1ug/ml and the plate was further incubated for one hour. Plates were then washed and finally 150ul of MSD GOLD read buffer B, containing ECL substrate was added and plates were placed in a MESO QuickPlex SQ 120 instrument (MSD, Rockville, MD, USA).

The MESO QuickPlex SQ 120 applies electric current to the wells and SUFO-TAG labelled anti human IgG emits ECL in the presence of ECL substrate, which is quantified by the instrument. Analysis was performed using MSD Discovery Workbench Software Version 4.0 which interpolates concentrations from the standard curves. Samples with ECL >2,000,000 units were considered to be above the range of detection and the plasma sample was repeated but with an initial, greater dilution of 1:20,000. If the ECL reading at the higher dilution remained >2,000000, the sample was repeated at a further higher dilution of 1:100,000.

To harmonise the assay output to the first World Health Organisation (WHO) international standard for anti-SARS-CoV-2 immunoglobulin (reported as international units (IU)/ml), eight serial dilutions (1:4) of the WHO reference serum (National Institute for Biological Standards and Control (NIBSC) code 20/136, National Institute for Biological Standards and Control, Potters Bar, Hertfordshire, UK) were run in duplicate on two occasions, and a conversion equation was derived from the linear curve of the standard curves for each of the four antibody targets to convert the ng/mL ECL reading to IU/ml, with the final results reported in IU/mL.

### Comparison with commercially-available quantitative and semi-quantitative assays

Reactivity against S1-RBD, spike and nucleocapsid was measured with the commercially available V-PLEX SARS-CoV-2 Panel 2 Kit (K15383U, MSD, Rockville, MA, USA), which uses ECL technology as outlined above but with antigens precoated to individual carbon spots rather than coupled with linkers. These kits comprise 96 well plates, precoated with antigens, proprietary blocker, diluent, wash buffer, detection antibody, read buffer, control sera and reference standard. Assays were performed as per manufacturer’s instructions. Briefly, precoated plates were blocked with 150ul of MSD blocker A per well for 30 minutes. Plasma samples were diluted 1 in 8,000 and 50ul was added to each well and incubated for 2 hours, then 50ul of SULFO-TAG anti human IgG was added and incubated for 1 hour. Plates were washed with MSD wash buffer between binding steps. 150ul of MSD gold read buffer was added and plates read with the MESO QuickPlex SQ 120 instrument, and analysed with MSD Discovery Workbench Software. Arbitrary Units interpolated from the reference standard were converted to IU/ml using a conversion factor provided by the manufacturer. Saturated plasma samples were repeated at a higher initial dilution of 1:40,000.

In addition, assay performance was compared with the Abbott SARS-CoV-2 IgG assay and the Abbott SARS-CoV-2 IgG II assay, chemiluminescent microparticle immunoassays (CMIA) (Abbott laboratories, IL, USA). The Abbott SARS-CoV-2 IgG is a semi-quantitative assay targeting the nucleocapsid protein and the Abbott SARS-CoV-2 IgG II is a quantitative assay targeting the RBD of the S1 subunit of the spike protein. Both assays are automated, two step immunoassays, run on the Architect i2000R platform (Abbott laboratories, IL, USA) and were performed as per the manufacturer’s instructions. Briefly SARS-CoV-2 antigen coated paramagnetic microparticles were incubated with plasma. Mixture was washed and anti-human IgG is added and incubated. After a further wash, a pre trigger and trigger solution was added and the resulting chemiluminescent reaction measured as a relative light unit. Sample to calibrator (S/C) signals of ≥1.4 and ≥50 were interpreted as reactive for the nucleocapsid and spike assays respectively.

### Statistical analysis

Continuous variables were summarised using median and interquartile range (IQR) and categorical variables as frequency and percent. Assay correlations were determined using Spearman’s rank correlation coefficient. For the sensitivity and specificity calculations, the limit of detection of the assay was set at 3 standard deviations above the mean signal of 38 plasma samples collected prior to the pandemic (pre-March 2020), while samples derived from subjects with COVID-19 confirmed by a positive SARS-CoV-2 PCR was considered as true positive samples. Receiver operating curves, area under the curve and Youden Index were constructed using the pROC package in R. The Youden Index is the point on the receiver operating curve where sensitivity plus specificity is maximal, and aims to select and optimal threshold value for a diagnostic test^13^. Kruskall-Wallis test was used to compare IgG levels between groups. All analyses were performed with R software, version 3.6.2.

## Results

### Patient demographics

A total of 193 samples from individuals with PCR confirmed COVID-19, 58 samples from vaccinated individuals and 52 samples from confirmed COVID-19 negative individuals were included in the analysis of the CEPHR COVID-19 Serology Assay, henceforth referred to as the convalescent group, vaccinated group and COVID negative group respectively (demographics are shown in Table 1). In the convalescent group, all individuals had acute onset of symptoms from March to May 2020 with median time from onset of acute symptoms to plasma collection being 99 days (IQR 35-182). Although most (68%) had mild initial disease by WHO severity^14^, subjects with a range of disease severity were included. Sequence of the infecting SARS-CoV-2 variant was available for 33 individuals from the convalescent group, with the pangolin lineages reflecting the predominant circulating variants in Ireland at the time of infection^15^, and no variants of concern were identified. In the vaccinated group, the majority (86%) had received 2 doses of BNT162b2. Although there were 23 (40%) within the vaccinated group who also had a documented history of PCR confirmed COVID-19, these were kept separate from the convalescent group for the sensitivity analysis. The COVID negative group consisted of 40 samples collected prior to July 2019 and 12 post-pandemic samples which had no history of COVID-19 and a PCR negative result prior to sampling.

**Table 1.** Participant Demographics

|  | Convalescent<br>(n =193) | Vaccinated<br>(n = 58) | COVID negative<br>(n = 52)* |
| --- | --- | --- | --- |
| Age (Median (IQR)) | 49 (40-63) | 46 (35 – 55) | 46 (39-53) |
| Female sex (n (%)) | 106 (55) | 32 (57) | 25 (48) |
| Caucasian ethnicity (n (%)) | 145 (75) | 42 (75) | 27 (52) |
| Days from symptom onset (Median (IQR)) | 99 (35-182) | - |  |
| Days from second vaccine (Median (IQR)) | - | 55 (36 – 104) |  |
| History of prior infection (n (%)) | 193 (100) | 23 (40) |  |
| Mild initial disease severity** (n (%)) | 131 (68) | 19 (82) |  |
| COVID-19 variant (pangolin lineage) (n = 33) (n (%)) |  |  |  |
| B.1.1 | 14 (42) |  |  |
| B.1.1.267 | 8 (24) |  |  |
| B | 3 (9) |  |  |
| Other (B.1, B.1.1.1, B.1.1.163, B.1.1.29, | 8 (24) |  |  |
| B.3, B.29, B.1.5,<br>B.1.1.10) |  |  |  |
| Vaccine (n (%)) | - |  |  |
| - BNT162b2 |  | 48 (86) |  |
| - mRNA-1273 |  | 7 (12) |  |
| - ChAdOx1-S |  | 1 (2) |  |

\*40 samples taken from subjects prior to the onset of the pandemic, 12 samples from individuals in early 2020 with confirmed negative PCR for SARS-CoV-2
\*\*By World Health Organisation severity grading<sup>14</sup>

### Assay performance

#### Assay parameters

The lower limit of detection of the assay was set using deidentified remnant plasma samples taken pre-pandemic and thus considered to be true negatives. Mean and standard deviation of the concentration of the pre-pandemic samples was calculated and lower limit of detection set at three standard deviations above the mean concentration, demonstrating significant differences between convalescent individuals and controls (p<0.001 for all antigens, Figure 1). The lower limit of detection of each antigen was 38 IU/ml for RBD, 27 IU/ml for S1, 18 IU/ml for S2 and 20 IU/ml for nucleocapsid. The upper limit of detection was determined by running serial dilutions of monoclonal antibody against each antigen, and set at the upper range of ECL (>2,000,000) above which the curve no longer demonstrated linearity (Figure 2).

**Figure 1.**
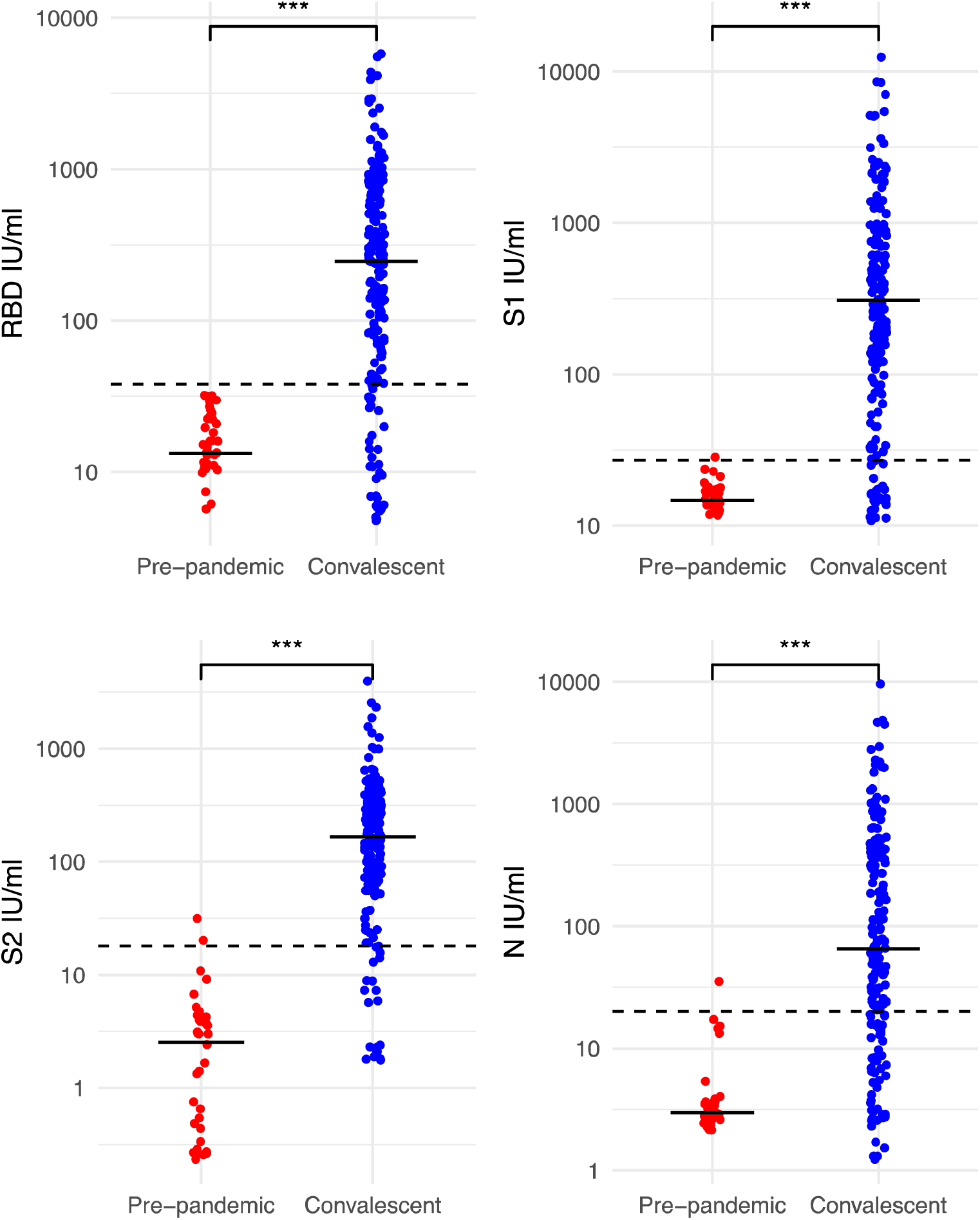
IgG levels in individuals with confirmed COVID-19 vs pre-pandemic controls Legend: IgG levels as measured by the CEPHR assay against 4 antigens, horizontal line represents median value in each group, dotted line indicates lower limit of detection. RBD – receptor binding domain, S1 – S1 subunit, S2 - S2 subunit, N - nucleocapsid. Significance testing performed with Kruskall Wallis tests, *** denotes p value <0.001.

**Figure 2.**
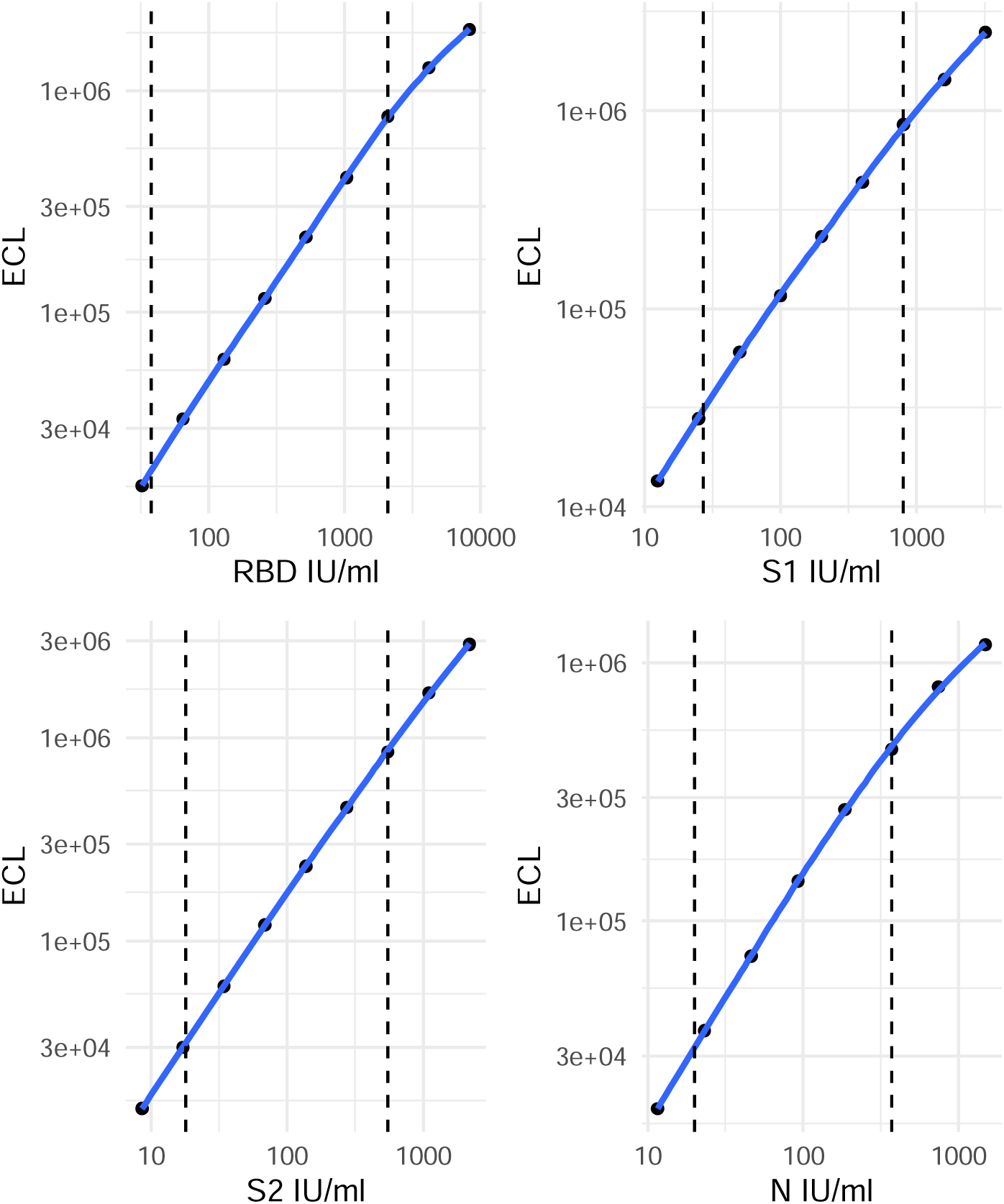
Standard curves for each component antigen in the CEPHR assay. Legend – Dashed vertical lines denote upper and lower limits of detection of the assay. ECL – electrochemiluminescence, RBD – receptor binding domain, S1 – S1 subunit, S2 - S2 subunit, N - nucleocapsid.

#### Assay precision

Intra-assay variability is the within run variation that represents the repeatability under the same conditions, whereas inter-assay variability (otherwise known as intermediate precision) represents the repeatability across different conditions^12^. The mean (standard deviation(SD)) intra-assay (within plate) coefficient of variation (CV) of 80 plasma samples run on the same plate was 3.9% (2.9%) for N, 3.8% (6.2%) for RBD, 3.8% (5.9%) for S1 and 3.9% (5.3%) for S2. The mean (SD) inter-assay CV derived from 5 samples run across 3 days by two different operators spanning a range of reactivity across the different antigens was 11% (6.5%) for N, 13% (5.7%) for RBD, 14% (8.9%) for S1 and 13% (5.1%) for S2 (Figure 3).

**Figure 3.**
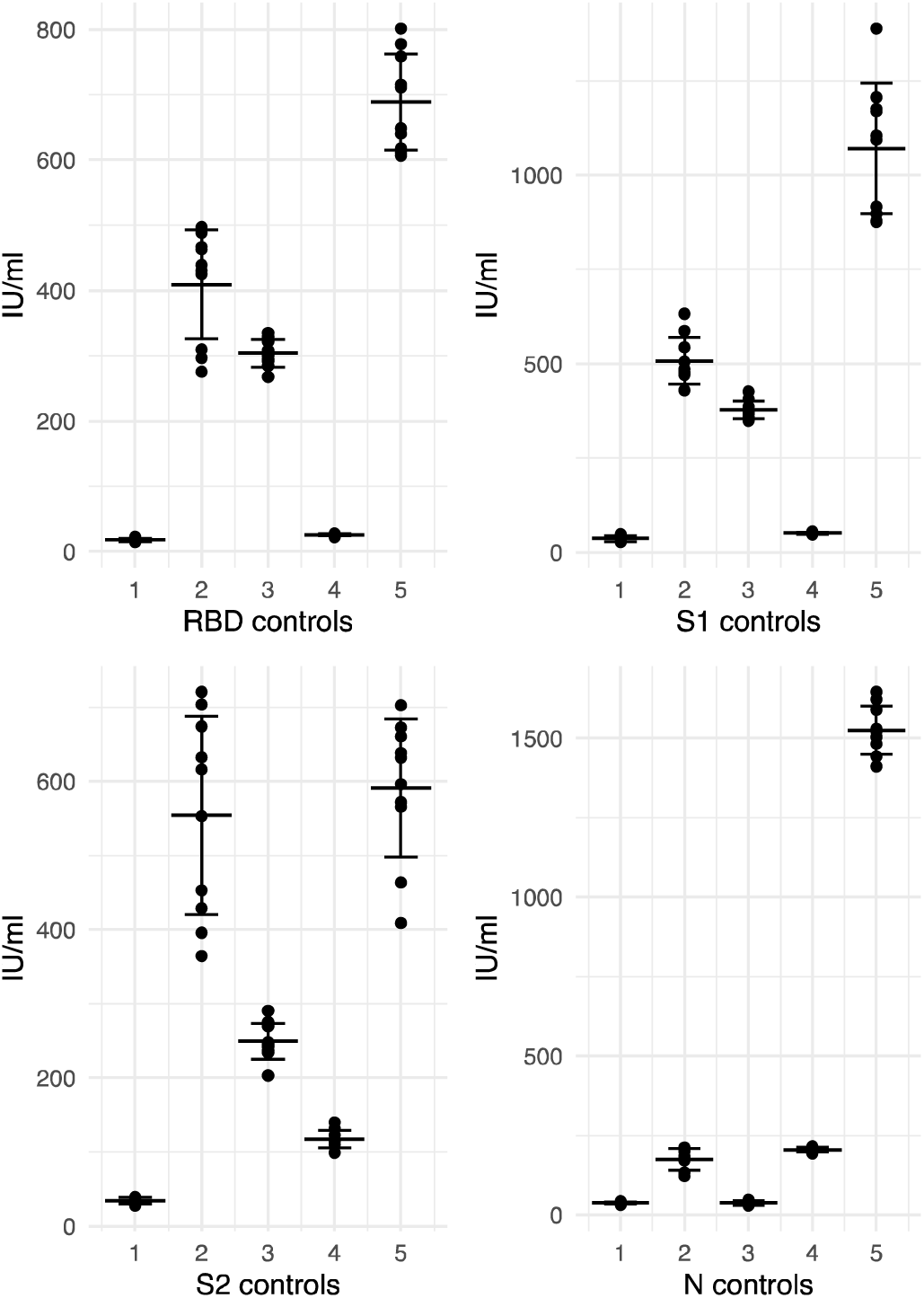
Antibody concentrations in 5 controls run on 10 plates across 3 days. Legend – Concentrations of each of the 5 controls run on 10 occasions across 3 days. Middle line represents the mean and upper and lower line represent one standard deviation from the mean.

#### Sensitivity and specificity

*I*n the convalescent group (n=193), overall sensitivity for each assay was; RBD 82% (95% confidence interval (CI) 76-87%), S1 86% (95% CI 81-91%), S2 88% (95% CI 83 – 92%) and N 72% (95% CI 64 – 78%). Sensitivity improved for the binding antibody targets when analysis included only individuals who were sampled more than 14 days from onset of symptoms (n=166), RBD 87% (95% CI 81 – 95%), S1 91% (95% CI 85 – 95%), S2 91% (95% CI 85 – 95%) but not for the N-target (73% (95% CI 66-80%)).

We explored specificity of the assay (to exclude COVID-19) in the COVID negative group (n = 52). Overall specificity was 96% (95% CI 87-99%) for RBD, 90% (95% CI 78-97%) for S1, 94% (95% CI 84-99%) for S2 and 90% (95% CI 78-97%) for N.

To further evaluate the performance of the assay we constructed receiver operating characteristic (ROC) curves for each antigen, using the 193 samples from the convalescent group as true positives and 38 pre-pandemic samples as true negatives respectively. The area under the curves (AUC) were 0.88, 0.91, 0.96 and 0.9 for RBD, S1, S2 and N respectively. To see if we could improve classification performance, we calculated the Youden index for each assay and recalculated sensitivity and specificity in the convalescent and COVID negative group, giving lower limits of detection of 31 IU/ml for RBD, 26 IU/ml for S1, 12 IU/ml for S2 and 3.4 IU/ml for N. Using these thresholds, overall sensitivity and specificity was 84% (95% 78 – 89%) and 96% (95% 87 – 99%) for RBD, 86% (95% 81 – 91%) and 88% (95% 76 – 96%) for S1, 91% (95% 86 – 95%) and 90% (95% 79 – 97%) for S2 and 89% (95% 84 – 93%) and 73% (95% 59 – 84%) for N, respectively. Although the lower limits of detection set by Youden Index gave the assay a higher sensitivity compared to the initially derived lower limits of detection, we determined that these thresholds did not improve overall assay performance given the corresponding decrease in specificity, and retained the initial thresholds for further analyses.

### Performance in Vaccinated individuals

In 58 samples from 56 vaccinated individuals, 100% (95% CI 94-100%) had both detectable RBD and S1 antibodies. In keeping with other reports, RBD and S1 titres were significantly higher in those with prior, documented SARS-CoV-2 infection (median (IQR) RBD 7,421 (3,909-21,462) IU/mL in those with previous infection versus 1,540 (811-3,209) IU/mL in those without prior infection, p<0.001). Within the vaccinated group, nucleocapsid antibody was detected in 20 (83%) of 24 individuals with a known history of COVID-19 and 6 (19%) of 32 individuals with no known history of COVID-19, giving an overall sensitivity of 83% (95% CI 63-95%) and specificity of 84% (95% CI 67%-95%), however given the potential for prior asymptomatic or pauci-symptomatic COVID-19, this may be an underestimate of the true specificity.

### Correlation with other immunoassays

Correlation coefficients comparing the CEPHR COVID-19 Serology Assay to two commercial assays are listed in table 2. Assays targeting the SARS-CoV-2 spike protein may target the full spike protein, S1 or S2 subunits, or the RBD, which lies within the S1 subunit. The CEPHR, MSD V-PLEX and Abbott assays all differ slightly in their target spike epitopes. The Abbott

**Table 2.** Correlation between assays

|  | Abbott Spike | Abbott N | V-PLEX S1 -<br>RBD | V-PLEX Spike | V-PLEX N |
| --- | --- | --- | --- | --- | --- |
| CEPHR RBD | 0.91 |  | 0.75 | 0.83 |  |
| CEPHR S1 | 0.9 |  | 0.73 | 0.81 |  |
| CEPHR S2 | 0.77 |  | 0.36 | 0.44 |  |
| CEPHR N |  | 0.74 |  |  | 0.81 |

Legend: Spearman's rho between different assays with similar antigenic targets. RBD – Receptor binding domain, S1 -Spike subunit 1, S2 - Spike subunit 2, N – Nucleocapsid. For all correlations $p < 0.001$ .

SARS-CoV-2 IgG II measures only anti-RBD IgG, the MSD V-PLEX measures both IgG against full spike and separately the S1-RBD, while the CEPHR COVID-19 Serology Assay distinguishes between RBD, S1 and S2 subunits, providing separate quantitative antibody readings against each of the three targets.

Using the Abbott SARS-CoV-2 IgG II, we measured 210 samples from across the three groups. There was excellent correlation between the Abbott IgG II and both CEPHR anti-RBD IgG (rho 0.91) and CEPHR anti-S1 IgG (rho 0.9, both p <0.001, Figure 5.), while CEPHR anti-S2 IgG was less tightly correlated (rho 0.77 p<0.001), in line with the difference in antigenic target.

In 188 samples measured on the MSD V-PLEX assay, counterintuitively, correlation was better between the V-PLEX full spike and both CEPHR RBD IgG (rho 0.83) and S1 IgG (rho 0.82, both p <0.001, Figure 4), than with the more antigenically similar V-PLEX S1-RBD and CEPHR RBD (rho 0.75) or S1 (rho 0.73, both p <0.001). Correlations with the CEPHR S2 were lower; V-PLEX S1-RBD and CEPHR S2 correlation was rho 0.36, p <0.001, while V-PLEX full spike and CEPHR S2 correlation was rho 0.44, p <0.001, with this weaker correlation again likely reflecting differences in antigenic target.

**Figure 4.**
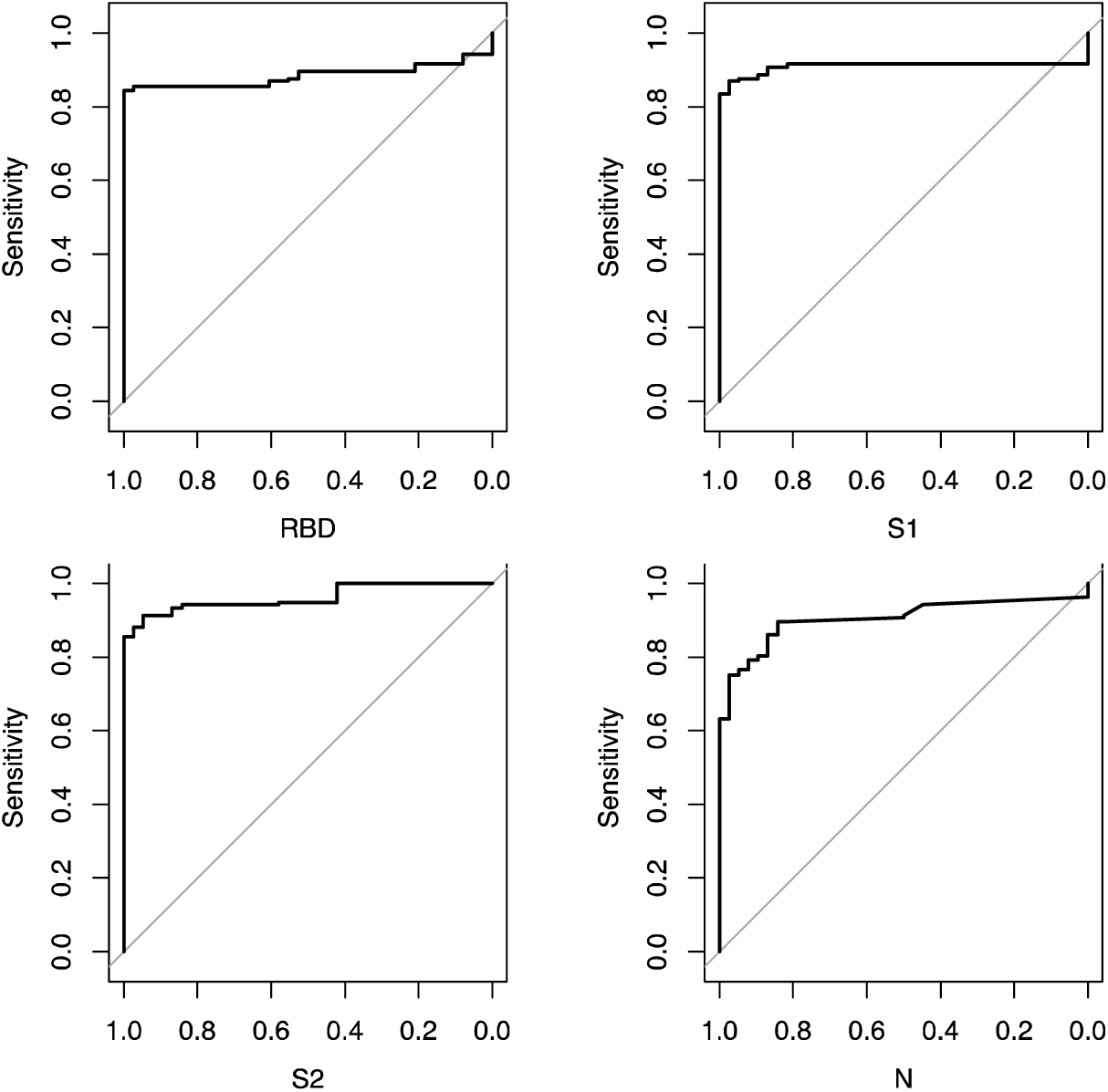
Receiver Operating Characteristic curves for each antigen. Legend: ROC curves constructed using the pROC package. Area Under the Curve (AUC) was 0.88 for RBD, 0.91 for S1, 0.96 for S2 and 0.9 for N respectively. RBD – receptor binding domain, S1 – S1 subunit, S2 - S2 subunit, N - nucleocapsid.

**Figure 5.**
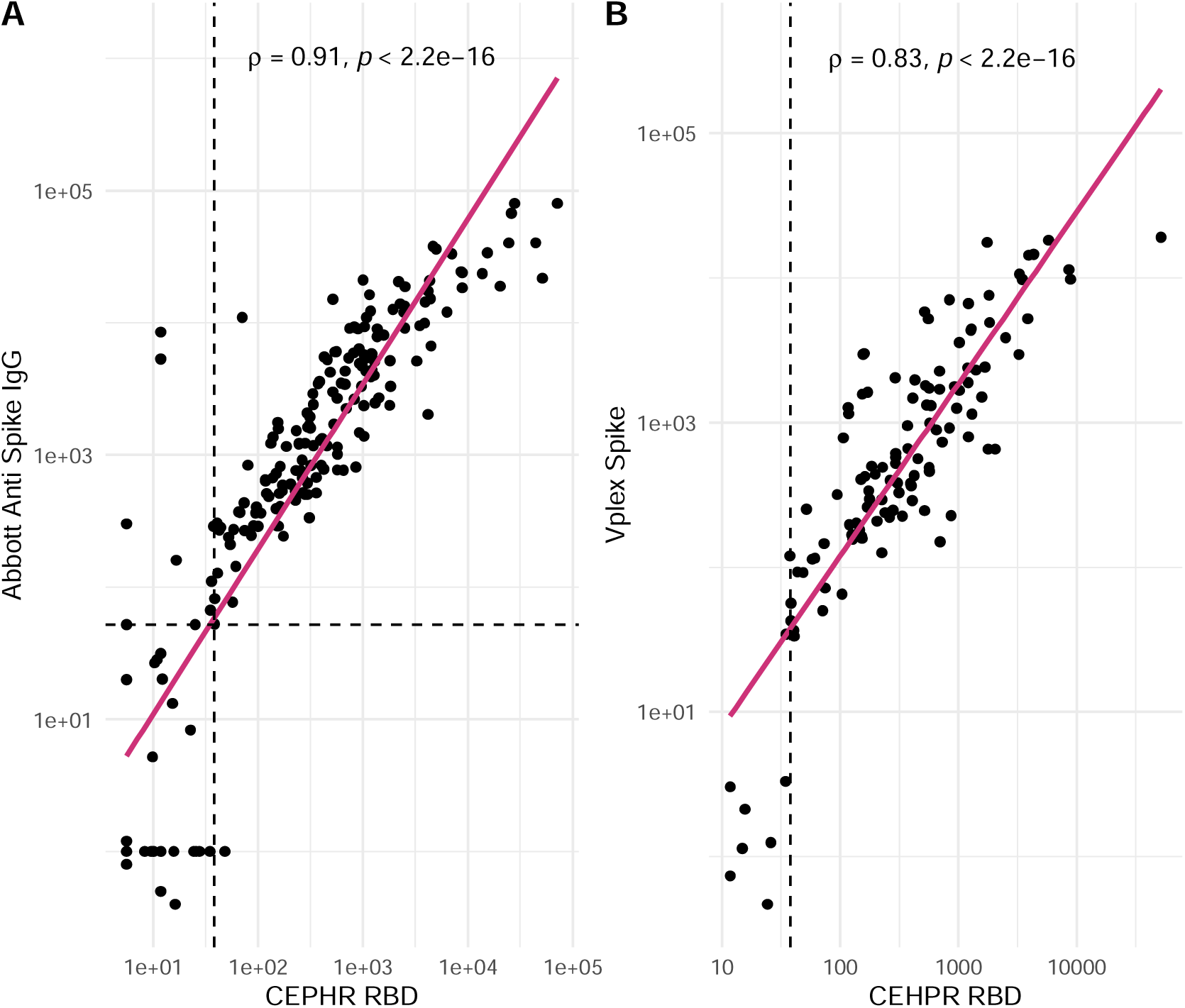
CEPHR RBD correlation with Abbott and MSD Vplex spike assays Legend: A: Correlation between CEPHR RBD and Abbott SARS-CoV-2 IgG II anti spike assay. Vertical dashed line represents CEPHR RBD positivity threshold, horizontal dashed line indicates Abbott positivity threshold. B: Correlation between CEPHR RBD and MSD V-PLEX Spike IgG. Vertical dashed line represents CEPHR RBD positivity threshold, no positivity threshold provided by MSD.

Correlation between the Abbott SARS-CoV-2 IgG assay, which targets the nucleocapsid protein, and the CEPHR N IgG was rho 0.75, p <0.001, while correlation between the MSD V-PLEX N IgG and CEPHR N IgG was 0.81, p<0.001.

## Discussion

This study describes the performance characteristics of the CEPHR COVID19 Serology Assay a new, quantitative, multiplex serologic assay for SARS-CoV-2 that is both sensitive and specific for detection of immunological responses to both COVID19 infection and SARS-CoV-2 vaccination. In addition the CEPHR COVID19 Serology Assay highly correlates with currently available commercial assays.

The CEPHR assay offers a number of advantages over the currently available commercial assays tested. The multiplex format allows for assessment of antibodies against multiple antigenic targets in one well, requiring a very small amount of plasma and optimising on both preparation time and reagents, while the 96 well plate format allows for high throughput testing. Results are normalised to WHO standardised IU/ml, which enables better quality control, not only between laboratories but between assays. Finally, the linker format permits future customisation, leaving the option to rapidly amend or add target antigens as new variants of SARS-CoV-2 emerge.

While overall sensitivity was high, and consistent with similar performance characteristics of commercial assays^16^, the different antigenic targets employed in the CEPHR COVID-19 Serology Assay displayed different performance characteristics. In particular, the CEPHR N IgG was the least sensitive overall, in line with the shorter reported half-life of anti-SARS-CoV-2 nucleocapsid antibodies. The half-life of nucleocapsid antibodies post infection has been shown to have a shorter than that of spike targeted antibodies^17^. Loss of nucleocapsid-specific IgG responses have been demonstrated in up to 35% of individuals, commonly less than 180 days from seroconversion.^18^

Multiple sources of evidence demonstrate positive correlation between antibody titres and protection against COVID-19^3,19^, however at an individual level the clinical relevance of a quantitative antibody result remains unclear^20^ due to the lack of a clear definition of a threshold of antibody response sufficient to provide protection, either against infection with SARS-CoV-2 or protection from development of severe disease if infection occurs. That currently available serologic assays use a wide variety of terminologies, techniques and reporting units further complicates progress towards identification of a meaningful threshold of immunity. In 2020, the World Health Organisation developed an international standard derived from pooled convalescent plasma to enable harmonisation of results across different assays and laboratories, and stressed the importance of its widespread adoption^21^. Despite this, many commercial quantitative serologic tests still report results in assay specific units, further complicating comparisons both between assays and between studies. Normalisation of the results of this assay to IU/ml, as has been performed with the CEPHR COVID19 Serology Assay permits comparison both between assays that use similar standardised units and between laboratories.

Although the CEPHR COVID19 Serology Assay has many advantages, it is not without limitations. The assay employs RBD derived from the Wuhan-Hu-1 reference strain of SARS-CoV-2. Although the test has been validated in convalescent plasma from individuals with confirmed SARS-CoV-2, these individuals were infected with the variants circulating in the first wave of infections in early 2020 and the performance of this assay against the different SARS-CoV-2 variants of concern (VOCs) that have emerged since is still under investigation. In addition, the vaccinated population was relatively small, with the majority of individuals less than 3 months from second dose vaccine. Given waning of post vaccine protection^22^, sensitivity of the assay may alter as time post vaccination increases. Additionally binding assays do not evaluate antibody function, such as neutralising capacity or antibody effector function, although these gold standard assays are time consuming and expensive and do not lend themselves to high throughput. However, correlation of the CEPHR COVID19 Serology Assay with these gold standard functional assays is ongoing.

Despite these limitations, the CEPHR SARS-CoV-2 Serology Assay is a robust, customisable, multiplex serologic assay for the detection of several different IgG specific to SARS-CoV-2, with multiple potential real world applications and performance characteristics that support its further development for use in both research and clinical settings.

## Data Availability

Access to pseudonymised data can be requested by submission to the All Ireland Infectious Diseases Cohort steering group, access will be granted depending on a data protection impact assessment and assessment of the research proposal.

## Funding

This work was supported by Science Foundation Ireland (grant numbers 20/COV/0305 and 20/COV/8549) and a philanthropic donation from Smurfit Kappa and the VACCELERATE the European Corona Vaccine Trial Accelerator Platform. GK is funded through a fellowship from the United States Embassy in Ireland.

## Acknowledgements

The authors wish to thank all study participants and their families for their participation and support in the conduct of the All Ireland Infectious Diseases Cohort Study.

The All Ireland Infectious Diseases Cohort Study Investigators:

Mater Misericordiae University Hospital: A. Cotter, E. Muldoon, G. Sheehan, T. McGinty, JS. Lambert, S. Green, K. Leamy. St Vincent’s University Hospital: G. Kenny, K. McCann, R. McCann, C. O’Broin, S. Waqas, S. Savinelli, E. Feeney, PWG. Mallon. CEPHR: A. Garcia Leon, S. Miles, D. Alalwan, R. Negi. Beaumont Hospital: E. de Barra, S. McConkey, K. Hurley, I. Sulaiman. University College Cork: M. Horgan, C. Sadlier, J. Eustace. University College Dublin: C. Kelly, T. Bracken. Sligo University Hospital: B. Whelan, Our Lady of Lourdes Hospital: J Low. Wexford General Hospital: O Yousif. University Hospital Galway: B. McNicholas. St Luke’s Hospital Kilkenny: G. Courtney. Children’s Health Ireland: P. Gavin.

## Notes

### Competing Interest Statement

Eoin Feeney has received consulting fees from Gilead, ViiV and Vidacare Ireland. Eoghan de Barra has received consulting fees from Sanofi Pasteur and honoraria/travel grant from Pfizer. Patrick Mallon has received honoraria and/or travel grants from Gilead Sciences, MSD, Bristol Myers Squibb, and ViiV Healthcare.

### Author Declarations

Office of the National Research Ethics Committee Ireland St. Vincents Healthcare Group, Ethics and Medical Research Committee and the Mater Misericordiae University Hospital gave ethical approval for this work

## References

1 Mallon PWG, Tinago W, Leon AG, et al. Dynamic Change and Clinical Relevance of Postinfectious SARS-CoV-2 Antibody Responses. Open Forum Infect Dis 2021; 8: ofab122.

2 Wei J, Stoesser N, Matthews PC, et al. Antibody responses to SARS-CoV-2 vaccines in 45,965 adults from the general population of the United Kingdom. Nat Microbiol 2021; 6: 1140–9.

3 Khoury DS, Cromer D, Reynaldi A, et al. Neutralizing antibody levels are highly predictive of immune protection from symptomatic SARS-CoV-2 infection. Nat Med 2021; 27: 1205– 11.

4 O’Brien MP, Forleo-Neto E, Musser BJ, et al. Subcutaneous REGEN-COV Antibody Combination to Prevent Covid-19. N Engl J Med 2021; 385: 1184–95.

5 Wheatley AK, Juno JA, Wang JJ, et al. Evolution of immune responses to SARS-CoV-2 in mild-moderate COVID-19. Nat Commun 2021; 12: 1162.

6 European Medicines Agency. COVID-19 vaccines. https://www.ema.europa.eu/en/human-regulatory/overview/public-health-threats/coronavirus-disease-covid-19/treatments-vaccines/covid-19-vaccines (accessed Feb 16, 2022).

7 Garcia-Beltran WF, Lam EC, Astudillo MG, et al. COVID-19-neutralizing antibodies predict disease severity and survival. Cell 2021; 184: 476–488.e11.

8 Plotkin SA. Correlates of Protection Induced by Vaccination. Clin Vaccine Immunol 2010; 17: 1055–65.

9 Kenny G, Mallon PWG. COVID19-clinical presentation and therapeutic considerations. Biochem Biophys Res Commun 2021; 538: 125–31.

10 Steensels D, Pierlet N, Penders J, Mesotten D, Heylen L. Comparison of SARS-CoV-2 Antibody Response Following Vaccination With BNT162b2 and mRNA-1273. JAMA 2021; 326: 1533.

11 US FDA. In Vitro Diagnostics EUAs - Serology and Other Adaptive Immune Response Tests for SARS-CoV-2. https://www.fda.gov/medical-devices/coronavirus-disease-2019-covid-19-emergency-use-authorizations-medical-devices/in-vitro-diagnostics-euas-serology-and-other-adaptive-immune-response-tests-sars-cov-2#isft3 (accessed March 10, 2022).

12 Marchese RD, Puchalski D, Miller P, et al. Optimization and validation of a multiplex, electrochemiluminescence-based detection assay for the quantitation of immunoglobulin G serotype-specific antipneumococcal antibodies in human serum. Clin Vaccine Immunol CVI 2009; 16: 387–96.

13 Fluss R, Faraggi D, Reiser B. Estimation of the Youden Index and its associated cutoff point. Biom J Biom Z 2005; 47: 458–72.

14 World Health Organisation. COVID-19 Clinical management Living guidance. 2021; published online Jan 25. https://www.who.int/publications/i/item/WHO-2019-nCoV-clinical-2021-1 (accessed Sept 23, 2021).

15 Mallon, PWG, Crispie F, Gonzalez G, et al. Whole-genome sequencing of SARS-CoV-2 in the Republic of Ireland during waves 1 and 2 of the pandemic. MedRxiv DOI:doi.org/10.1101/2021.02.09.21251402;

16 Patel EU, Bloch EM, Clarke W, et al. Comparative Performance of Five Commercially Available Serologic Assays To Detect Antibodies to SARS-CoV-2 and Identify Individuals with High Neutralizing Titers. J Clin Microbiol 2021; 59. DOI:10.1128/JCM.02257-20.

17 Cohen KW, Linderman SL, Moodie Z, et al. Longitudinal analysis shows durable and broad immune memory after SARS-CoV-2 infection with persisting antibody responses and memory B and T cells. Cell Rep Med 2021; 2: 100354.

18 Krutikov M, Palmer T, Tut G, et al. Prevalence and duration of detectable SARS-CoV-2 nucleocapsid antibodies in staff and residents of long-term care facilities over the first year of the pandemic (VIVALDI study): prospective cohort study in England. Lancet Healthy Longev 2022; 3: e13–21.

19 Feng S, Phillips DJ, White T, et al. Correlates of protection against symptomatic and asymptomatic SARS-CoV-2 infection. Nat Med 2021; 27: 2032–40.

20 U.S. Food and Drug Administration. Antibody Testing Is Not Currently Recommended to Assess Immunity After COVID-19 Vaccination: FDA Safety Communication. https://www.fda.gov/medical-devices/safety-communications/antibody-testing-not-currently-recommended-assess-immunity-after-covid-19-vaccination-fda-safety.

21 Knezevic I, Mattiuzzo G, Page M, et al. WHO International Standard for evaluation of the antibody response to COVID-19 vaccines: call for urgent action by the scientific community. Lancet Microbe 2021; : S2666524721002664.

22 Israel A, Shenhar Y, Green I, et al. Large-scale study of antibody titer decay following BNT162b2 mRNA vaccine or SARS-CoV-2 infection. MedRxiv Prepr Serv Health Sci 2021; : 2021.08.19.21262111.

